# Experiences of Communities with Lebanon’s Model of Care for Non-Communicable Diseases (NCD): A Cross-Sectional Household Survey from Greater Beirut

**DOI:** 10.1101/2022.11.24.22282716

**Authors:** Ibrahim R. Bou-Orm, Pol deVos, Karin Diaconu

**Author notes:** **Corresponding author:** Ibrahim R. Bou-Orm; NIHR Global Health Research Unit on Health in Situations of Fragility; Institute for Global Health and Development; Queen Margaret University, Edinburgh; EH21 6UU; United Kingdom.

## Abstract

**Introduction:** Health systems in fragile settings face multiple challenges in the implementation of responsive Non-Communicable Disease (NCD) care models. Models based on comprehensive person-centred primary care approaches can improve health system responsiveness and trust in healthcare. In Lebanon, NCDs dominate the health profile, but the health system is fragmented with evidence suggesting varied experiences with the care model. This study aims to identify people’s perceptions of the Lebanese care model for NCDs and trust in the health system among others, and test association between them.

**Methods:** This study is a household survey using multistage random sampling and targeting adult community members (both Syrian and Lebanese) living with NCDs in Greater Beirut. Three main outcomes (barriers to care seeking, perceptions of the care model and trust in healthcare) were assessed including by multiple linear regressions.

**Results:** A total of 941 respondents participated in this study. Reported NCDs were hypertension (51.3%) and diabetes (34.5%), followed by chronic respiratory conditions (21.9%) and other cardiovascular diseases (20.0%). Communities reported seeking care from different sources. While 78% of Lebanese participants had visited private clinics at least once within the 6 months preceding the survey, 56% of Syrian refugees had done so. Determinants of access to care were health coverage, gender, and employment among Lebanese, and socio-economic status among Syrian refugees. Lebanese community members had more positive perceptions of the care model compared to Syrian refugees and determinants included socio-demographic characteristics and the type of providers. Trust in the health system was higher among Syrian compared to Lebanese participants and was significantly influenced by the care model score and barriers to care seeking.

**Conclusion:** Our study generated evidence about the experience of people living with NCDs with Lebanon’s care model, and can inform service delivery reforms towards a more inclusive person-centred approach.

- What is already known on this topic
  - *Health systems in fragile settings struggle to implement responsive care models for NCDs with limited quantitative evidence exploring community perceptions of care models*.
- What this study adds
  - *Lebanese and Syrian communities living with NCDs in Greater Beirut experience challenges in access to care along with gaps in the continuity and comprehensiveness of services affecting trust in the health system*.
  - *Inequities based on socio-economic characteristics exist with vulnerable groups being more affected by barriers to care and negative experiences with services*.
- How this study might affect research, practice or policy
  - *This study identifies reform opportunities of Lebanon’s care model for NCDs and provides a baseline assessment of the care model*.

## Background

Non-communicable diseases (NCDs) are one of the most pressing health challenges of our time. Including cardiovascular diseases, cancers and mental health disorders among others, NCDs make up 63.8% of the global disease burden and are the primary cause of disability and mortality globally.^1^ They additionally present substantive risks when occurring in conjunction with other conditions, as also evident from the COVID-19 pandemic.^2^

Addressing the NCD burden is of utmost priority and potential solutions encompass whole of government and population-based approaches which can assist with both primary and secondary prevention, such as the WHO Best Buys, as well as implementation of highquality, comprehensive, and continuous care models.^3^ Given the high global and local disease burden in many countries, such a model can be considered the cornerstone of a health systems’ service delivery. Indeed, the 2008 WHO report on Primary Health Care (PHC) considered the implementation of a comprehensive person-centred primary care model as a key determinant for improving health system responsiveness to population health needs, including of those affected by NCDs.^4^

Health systems in situations of fragility ^5,6^ face multiple but unique challenges in relation to the implementation of NCD care models. First, in many countries that have faced conflict or major crises such as environmental shocks, both donor aid and service delivery priorities have historically focused on sexual and reproductive health, child health, and on addressing a high burden of injuries. ^7^ While priorities have evolved and now include provision for mental health and psychosocial support, comprehensive support for NCDs in situations of crisis is still rare despite high population prevalence.^7,8^ Second, for countries which are undergoing substantive socio-political unrest, but also economic challenges, NCD service delivery may be particularly needed but costly to set up.^9^ For example, skilling up existing health workers with limited NCD experience,^10^ investing in the continued supply and maintenance of medical devices and medications needed for diagnosis and disease management,^11^ as well as adapting existing, or creating new, health management and information systems^12^ require relatively high up-front costs, despite likely cost-effectiveness of such interventions.^13^ Across all these situations, successful implementation of a person-centred comprehensive and continuous care model is particularly difficult.

Where such care models are absent or ill-implemented, as they are in many situations of fragility, the trust of communities and service users in the health system may be affected.^14^ Fragility can also arise at the interface between community and health systems and manifest in this loss of trust.^6^ For example, trust in the health system could affect access to and use of medical care as well as affect a persons’ relationship with providers.^15^ This relationship is essential in the case of NCDs given the prolonged nature of service utilization that is needed to maintain disease control and prevent further complications. Despite its importance, trust in health systems is understudied, especially in low- and middle-income settings. ^15^

This paper focuses on the case of Lebanon, an extremely fragile setting, and the perceptions of persons affected by common NCDs regarding the care model they encounter at health facilities. In Lebanon, NCDs dominate the population health profile but care for the conditions is affected by the general fragmentation of health services.^16^ A system analysis of NCD prevention and control in Lebanon suggested that persons of diverse socio-economic backgrounds, and under different coverage schemes, experience different challenges in access to healthcare and that health seeking behaviours of affected persons are diverse but critically shaped by the role of trust in the overarching health system.^17,18^ The current study builds on these insights and seeks to identify people’s perceptions of the Lebanese care model for NCDs and trust in the health system among others, and test association between them.

## Methods

### Design, aims and objectives

This cross-sectional study aims to survey the perceptions of adult community members (Syrian refugee or Lebanese host community members) living with NCDs in the Greater Beirut area in relation to the healthcare model they encounter when seeking care for their condition.

The specific objectives of the study were to:

1. describe how persons access care: from whom, what barriers are encountered, what factors influence these barriers;
2. describe the perceptions of NCD patients in Greater Beirut in relation to the care model they encounter, including examining differences between Lebanese and Syrian participants;
3. assess the levels of trust of the aforementioned participants in the health system and identify whether this is associated with self-reported health status.

### Sampling

Multistage random cluster sampling was used within each sub-district of the Greater Beirut area, using the Probability Proportional to Size (PPS) approach and based on an existing sampling frame used in previous surveys.^19^ Within clusters, households were randomly selected and one eligible participant was recruited per household until the final sample size of 384 Syrian participants and 576 Lebanese participants was reached (see Appendix 1 for sample size estimation).

### Participant recruitment

As data collection was due to take place during the height of the COVID-19 pandemic, researchers employed a data collection agency offering phone-based data collection services. The agency already had contact details of community members agreeing to take part in health-related research within the targeted geographical location. Potential participants were called via telephone, an information sheet and oral consent form were shared, and participants were then asked if they can be contacted after 2 hours or at a different time to check their willingness to participate. Data collection was conducted between October and December 2020.

### Data collection

Data was collected in Arabic, using a newly developed questionnaire (see Appendix 2) which covered the following variables and scales:

- Care-seeking practices over the last 6 months: number of consultations or visits to health facilities and providers, access to NCD care, affordability of NCD care and out of pocket expenditures.
- Care model characteristics including regular and trusted relationship, continuity of care, comprehensiveness, coordination and other potential attributes from the 2008 WHO report on primary health care,^4^ and the Johns Hopkins Primary Care Assessment tool.^20^ Perceptions on these care model characteristics were gathered in order to be aggregated into a care model score – the higher the score, the more positive the view of persons with NCDs regarding the care they receive.
- Trust with 8 domains (honesty, communication, confidence, competence, confidentiality, fairness, fidelity and systems trust) as from Ozawa & Sripad (2013). ^15^
- Others: socio-demographic characteristics including age, gender, health coverage, and social capital (structural dimension only),^21^ as well as self-reported overall health status.^22^

The survey was piloted on 30 participants who met the eligibility criteria prior to the implementation of the full survey. Few edits to the Arabic version were made for better understanding and one item was removed from the scale on the features of the care model as respondents found it similar to another item. Examples were added to questions flagged to be difficult by data collectors in order to avoid any misinterpretation of questions and the risk of providing different clarifications by different data collectors during the full execution of the survey (to avoid information bias).

### Data analysis

The data was cleaned and analysed for all participants, as well as by sub-group, distinguishing between Lebanese and Syrian participants. Descriptive analyses were conducted in relation to each variable and also sample characteristics. Continuous variables were reported using means and standard deviations whereas categorical variables were tabulated and reported using counts and proportions.

Bivariate analyses assessed 1) the differences in sources of health seeking and health status by sub-group, and 2) the associations between selected socio-demographic characteristics and health-related variables with three main outcomes. The latter were indices and total scores referring reflective of barriers to care seeking, perceptions of the care model, and trust in the health system (see Appendix 2 for details on which questions were used as basis for calculation). Statistical tests included: t-tests; ANOVA or Kruskal-Wallis; Pearson’s correlation test; and Chi-square tests. For each outcome, three multiple linear regressions (one for each subgroup according to nationality and one for the whole sample) were developed including all variables which were significantly associated with the outcome in the bivariate analysis (p-value < or = 0.05), in order to determine their joint effects. Statistical analyses were conducted using SPSS v.23.

### Patient and Public Involvement

Patients or the public were not involved in the design, or conduct, or reporting, or dissemination plans of our research. However, a previous qualitative and participatory research exploring the dynamics of NCD control in Greater Beirut among communities informed the design and selection of outcomes of this study.

### Ethical considerations

Ethical approval was ensured from the QMU Research Ethics Panel and the ethics committee of the Saint-Joseph University of Beirut (USJ).

### Findings

We first offer an overview of sample characteristics and then present the three main outcomes in three sections.

### Sample characteristics

A total of 941 participants (574 Lebanese and 367 Syrian) were recruited to the study. Participant characteristics are summarised in Table 2. Overall, the majority of participants were between 46-55 years and male. Most participants were married (76%) and from a low educational background: 56.2% with only primary or secondary school attainment, with an unequal distribution by nationality (37.8% among Lebanese vs 88% among Syrian refugees). 52% and 50% of the group were unemployed and of low-socio-economic status (SES) respectively. In terms of health coverage, only 36% of the Syrian refugee community and about 67% of the Lebanese community reported having a formal health coverage.

**Table 1:** Eligibility criteria of participants.

| **Eligibility** | **Non-eligibility** |
| --- | --- |
| Nationality (Syrian or Lebanese) | Nationality (any other than Syrian or Lebanese) |
| Residing in the Greater Beirut area | Not residing in Greater Beirut |
| NCD officially diagnosed OR on treatment for one of the four common non-communicable chronic diseases: cardiovascular diseases, chronic respiratory diseases, diabetes and cancer. | Individual is healthy and not taking treatments OR (NCD not officially diagnosed AND no NCD treatment taken) |

**Table 2.** Socio-demographic characteristics of participants by nationality (N=941)

| **Variables** | **Lebanese**<br>**(n=574)** | **Syrian**<br>**(n=367)** | **Total**<br>**(n=941)** |
| --- | --- | --- | --- |
| **Age categories (n, %)** |  |  |  |
| Less than 25 | 57 (9.9) | 33 (9.0) | 90 (9.6) |
| 26 – 35 | 51 (8.9) | 51 (13.9) | 102 (10.8) |
| 36 – 45 | 75 (13.1) | 101 (27.5) | 176 (18.7) |
| 46 – 55 | 159 (27.7) | 102 (27.8) | 261 (27.7) |
| 56 – 65 | 130 (22.6) | 47 (12.8) | 177 (18.8) |
| 66 – 75 | 72 (12.5) | 29 (7.9) | 101 (10.7) |
| More than 75 | 30 (5.2) | 4 (1.1) | 34 (3.6) |
| **Age in years (mean; SD)** | 51.0 (15.9) | 45.6 (13.6) | 48.9 (15.2) |
| **Gender (n, %)** |  |  |  |
| Male | 301 (52.4) | 199 (54.2) | 500 (53.1) |
| Female | 273 (47.6) | 167 (45.5) | 440 (46.8) |
| Prefer not to mention | 0 (0.0) | 1 (0.3) | 1 (0.1) |
| **Marital status (n, %)** |  |  |  |
| Married | 417 (72.6) | 298 (81.2) | 715 (76.0) |
| Widowed | 43 (7.5) | 31 (8.4) | 74 (7.9) |
| Single | 107 (18.6) | 35 (9.5) | 142 (15.1) |
| Separated/Divorced | 7 (1.2) | 3 (0.8) | 10 (1.1) |
| **Educational level (n, %)** |  |  |  |
| Primary school or less | 55 (9.6) | 188 (51.2) | 243 (25.8) |
| Secondary school | 162 (28.2) | 124 (33.8) | 286 (30.4) |
| High school | 191 (33.3) | 37 (10.1) | 228 (24.2) |
| University | 166 (28.9) | 18 (4.9) | 184 (19.6) |
| **Employment status (n, %)** |  |  |  |
| Employed | 266 (46.3) | 153 (41.7) | 419 (44.5) |
| Unemployed | 278 (48.4) | 211 (57.5) | 489 (52.0) |
| Retired | 30 (5.2) | 3 (0.8) | 33 (3.5) |
| **Crowding index (mean ± SD)** | 1.23 (0.52) | 2.6 (1.22) | 1.78 ± 1.10 |

| **Socio-economic status (SES) (n, %)** |  |  |  |
| --- | --- | --- | --- |
| Low SES | 166 (28.9) | 306 (83.4) | 472 (50.2) |
| Middle SES | 293 (51.0) | 43 (11.7) | 336 (35.7) |
| High SES | 115 (20.0) | 18 (4.9) | 133 (14.1) |
| **Health coverage (n, %)** |  |  |  |
| Yes | 387 (67.4) | 132 (36.0) | 519 (55.2) |
| No | 187 (32.6) | 235 (64.0) | 422 (44.8) |

\* 19 participants were excluded as parents reported information on non-adult patients living with NCD

Reported NCDs in our sample were hypertension (51.3%) and diabetes (34.5%), followed by chronic respiratory conditions (21.9%) and other cardiovascular diseases (20.0%) (see Figure 1).

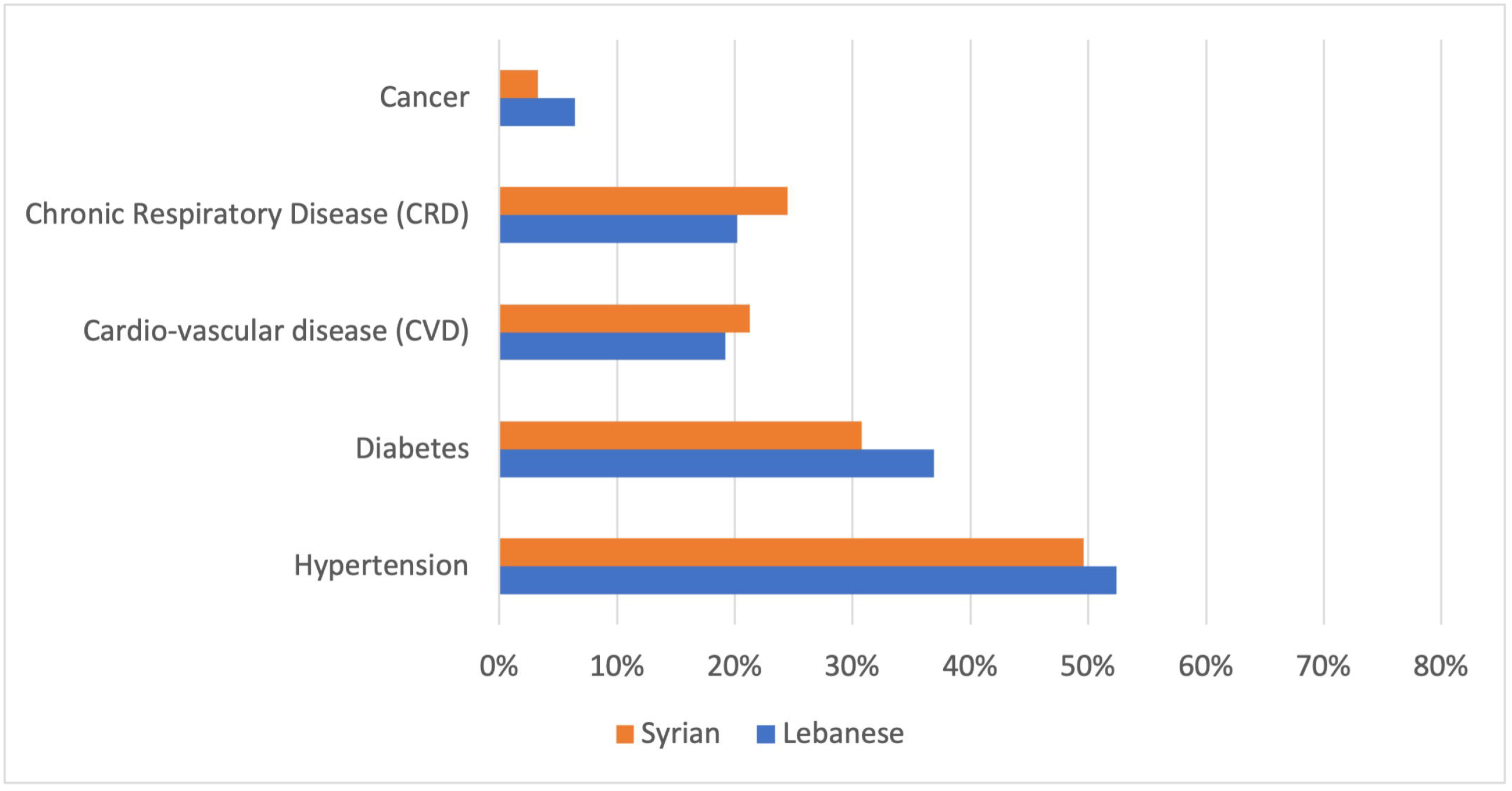

### Section 1: Access to care

#### Where health care was sought

The two studied communities reported seeking care from different sources. While 78% of Lebanese participants had visited private clinics at least once within the 6 months preceding the survey, 56% of Syrian refugees had done so. 21% of Syrian patients reported only one visit to private clinics.

The percentages of visits to primary care centres and dispensaries differ among groups as well: 40% of Lebanese respondents had visited a primary care centre/dispensary at least once in the same period compared to 68% of Syrian respondents.

The pharmacy was identified as a source of non-physician consultations for both communities with higher demand among Lebanese participants (56% reporting at least one consultation visit – compared to 40% among Syrian refugees).

About one fifth of the sample reported at least one visit to the emergency department of local hospitals for NCD-related complaints (24% among Lebanese vs 15% among Syrian respondents) and about 17% of all respondents were admitted to the hospital during the 6 months preceding the survey (21% among Lebanese compared to 11% among Syrian refugees).

#### Support for health seeking

When asked about the types of persons they would approach for support in case of urgent hospitalization, most Lebanese participants (92.5%) reported the ability and willingness to contact their immediate family and relatives, compared to 77.4% of Syrian participants. Other sources named by Syrian refugees were: NGOs and UN agencies (8.2%), neighbours (5.2%), and friends (4.9%).

#### Self-reported health status

About 55% of Lebanese participants reported a good or very good status – compared to only 39% among Syrian refugees living with NCD (see Figure 2).

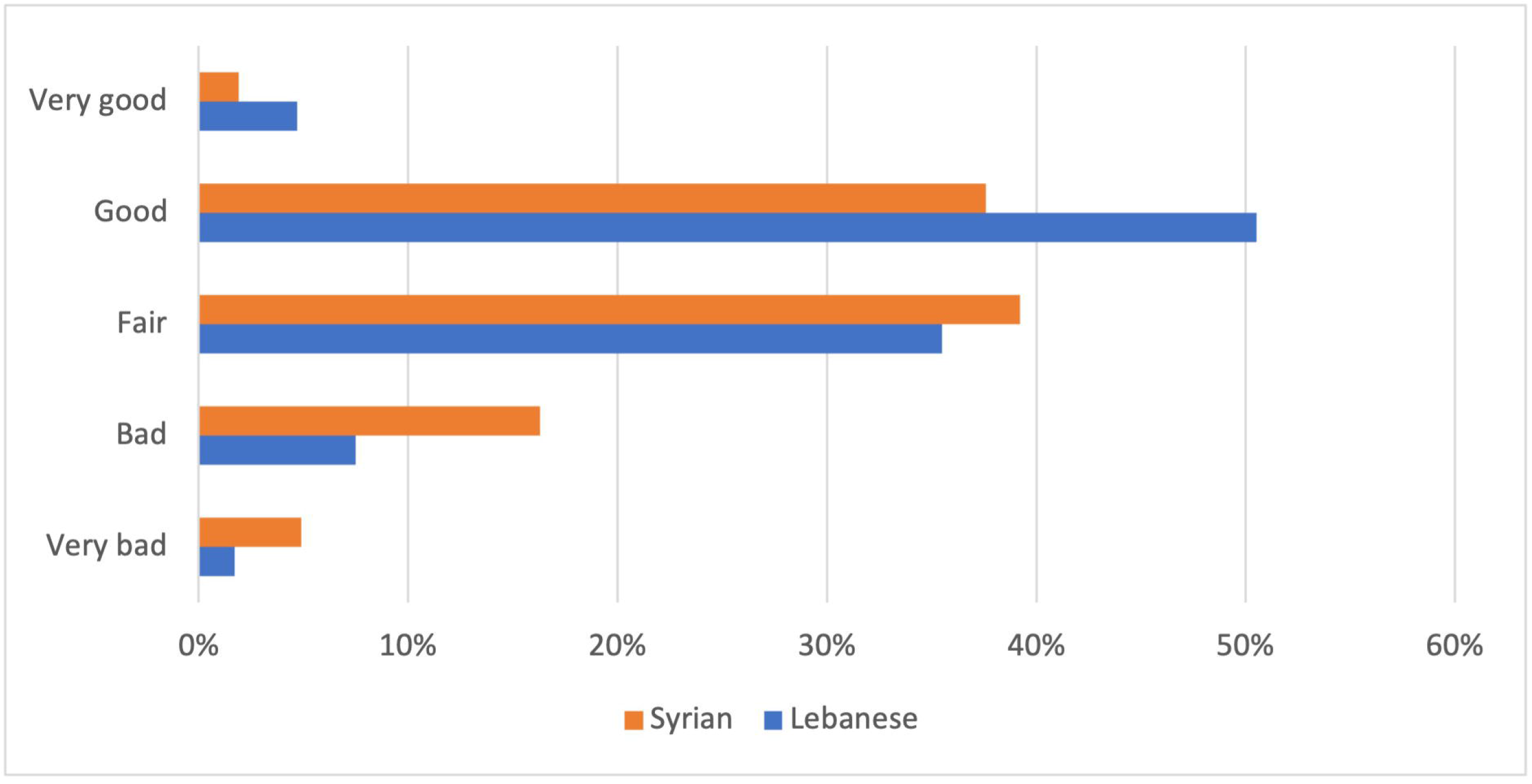

#### Barriers to care-seeking

The primary barrier to care seeking was the same for both communities, and identified as financial barriers; the least relevant barrier was noted as cultural. All barriers scored higher among the Syrian refugee community compared to the Lebanese community with the widest difference at the level of financial affordability of healthcare (mean difference of 1.06 over 5). All differences were statistically significant except for psychological barriers to healthcare. Appendix 3 provides a different presentation of findings by categories of answers. For instance, while about 21% of the Lebanese sample reported encountering financial barriers to care often or every time they seek healthcare, this proportion increases significantly to 62% among the Syrian refugee group.

As we anticipated that financial issues would present a main barrier to health seeking, health expenditures as percentage of monthly income were also assessed. Major differences within and between the study groups exist with higher percentages among Syrian refugees as about half of Syrian respondents pay at least 20% of their monthly income on health expenditures, compared to 27.1% among Lebanese respondents (see Appendix 4).

#### Influences on barriers to care-seeking

When considering the total impact of barriers on care-seeking, the total barriers to care seeking score suggests that Syrian refugees experience more barriers than host communities (Figure 3). However, when considering the median barrier score, responses from the two communities appear similar. Bivariate analyses suggest that lower socioeconomic status, unemployment, lower educational background, being a woman and the absence of health coverage all have significant bearing on whether persons experience barriers to care seeking (data not shown).

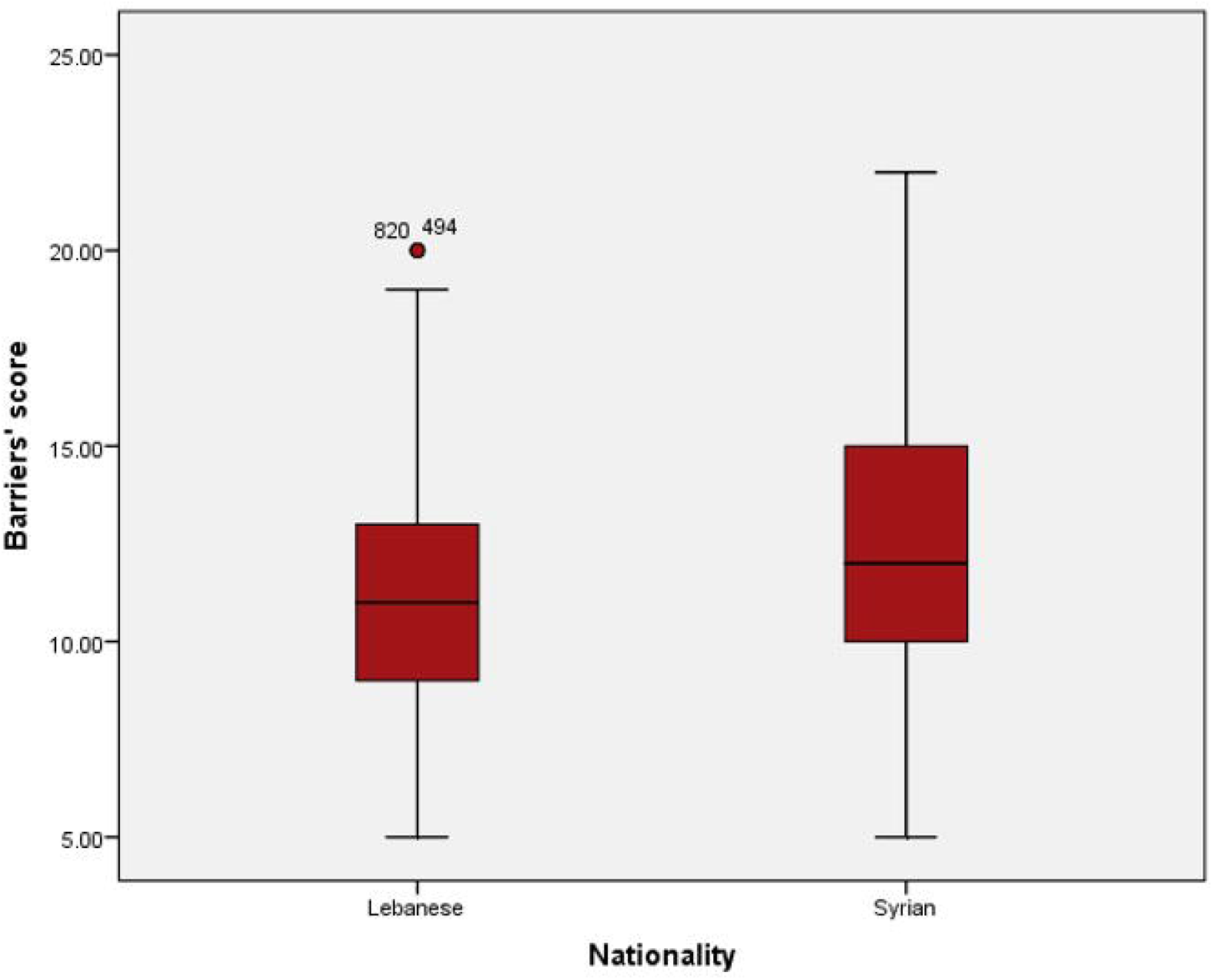

Table 3 offers an overview of the multivariable linear regression models of barriers across all participants and for the subgroups. Among Lebanese participants, the presence of health coverage, male gender, and being in employment were the three variables most strongly associated with a decrease in the barrier’s score. Socio-economic status also has bearing, with higher socio-economic status being associated with an estimated decrease of 0.55 (95%CI = 0.18 – 0.92) for each level of SES as well as higher education with an estimated decrease of 0.38 (95%CI = 0.05 – 0.71). In contrast, being female and additional years of age are associated with increases in the barriers’ score. For Syrian refugees, only being in employment and higher SES were still significantly associated with decreases in the barriers score. No other variables were significantly associated with the total score.

**Table 3.** Multivariable linear regressions of barriers to care (outcome) and covariates among people living with NCDs in Greater Beirut – Lebanon (2021)

| Variables | Multivariable models |  |  |
| --- | --- | --- | --- |
|  | Lebanese (n=574) | Syrian (n=367) | All (N=941) |
|  | Estimate [95% CI] | Estimate [95%CI] | Estimate [95% CI] |
| Age | **0.04 [0.02; 0.06]** | 0.02 [-0.03; 0.05] | **0.03 [0.01; 0.05]** |
| Gender (baseline = male) | **0.74 [0.19; 1.29]** | 0.25 [-0.57; 1.06] | **0.62 [0.17; 1.07]** |
| Marital status | 0.14 [-0.23; 0.51] | - 0.42 [-0.93; -0.08] | - 0.10 [- 0.39; 0.19] |
| Education | **- 0.38 [-0.71; -0.05]** | -0.23 [-0.65; 0.18] | **- 0.36 [- 0.62; - 0.10]** |
| Current employment | - 0.66 [-1.25; -0.06] | - 1.48 [-2.36; -0.60] | - 0.92 [- 1.41; - 0.44] |
| Socio-economic status | - 0.55 [-0.92; -0.18] | - 1.12 [-1.74; -0.50] | - 0.71 [- 1.02; - 0.40] |
| Nationality (baseline = Lebanese) | NA | NA | **0.87 [0.32; 1.43]** |
| Health coverage | **-1.35 [-1.89; -0.80]** | - 0.63 [-1.30; 0.04] | **- 1.05 [- 1.48; - 0.63]** |
| Willingness to approach immediate family/relatives for help | NS | - 0.44 [-1.17; 0.29] | - 0.16 [- 0.75; 0.42] |

### Section 2: Perceptions on the NCD care model

In this section, we summarize the perceptions of communities regarding the NCD care model. Perceptions are summarised according to the features of a person-centred primary health care model (for full list of questions that survey participants answered relating to this see Appendix 2). For full findings of the analyses please see Appendix 5. The higher the care model score, the better the perceptions of surveyed participants regarding the care they receive; perceptions are summarised according to domains below.

#### Availability of a regular and trusted health provider

At least 83% of the Lebanese participants either agreed or strongly agreed with each of the statements about having a stable and regular relationship with an accessible main provider as an entry point to the health system. This percentage is 62% among the Syrian refugee group.

#### Continuity of care

About 80% of Lebanese agreed or strongly agreed with being able to regularly use health services to follow-up on their condition(s). Only 49% of Syrian refugees said this.

#### Comprehensiveness of services

When asked about care coordination, the coordination of information and of services between different providers were acknowledged positively by about 85% and 64% of Lebanese and Syrian respondents respectively.

Only 78% of Lebanese respondents agreed they received appropriate care for all their health problems, and a lower percentage (68%) agreed that they can access secondary and tertiary NCD prevention services such as cancer screening and early detection of NCD complications. In contrast, among Syrian respondents, 46% agreed they received appropriate care, and 34% said they could access relevant secondary and tertiary prevention services.

#### Person-centredness

Only 67% of the Lebanese, and 43% of the Syrian participants, acknowledged that their health providers know them very well as persons and not just their medical condition(s). A higher percentage (81% for Lebanese and 65% for Syrian) acknowledged they were able to share their opinion about the provided care and get explanations from health providers.

Finally, at least 87% of respondents in both communities agreed with statements on the cultural competence of providers. There were no major differences between the two communities.

#### Influences on overall perceptions of the care model

While outliers exist across both groups, Lebanese respondents had significantly better perceptions of the care model compared to Syrian respondents (Figure 4).

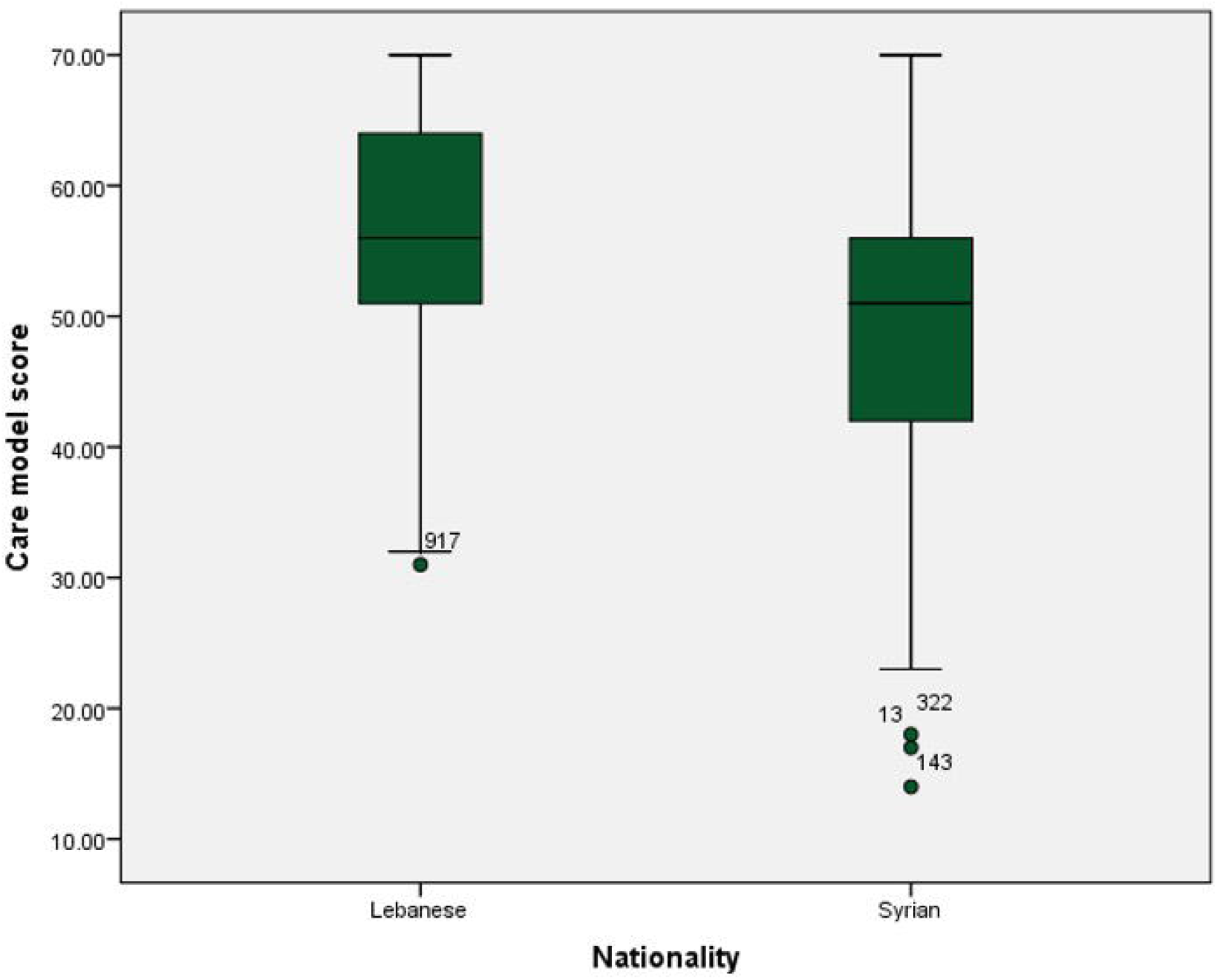

Bivariate and multivariate analyses investigated the association between the total care model score and socio-demographic characteristics of participants as well as barriers to care, type of main provider and sources of healthcare seeking (e.g. visits to PHC centres). Appendix 6 provides the detailed results of bivariate analyses, and regressions are presented in Table 4.

**Table 4.** Multivariable linear regressions of care model score (outcome) and other covariates among people living with NCDs in Greater Beirut – Lebanon (2021)

| **Variables** | **Lebanese (n=574)**<br>Estimate [95% CI] | **Syrian (n=367)**<br>Estimate [95% CI] | **Both communities**<br>**(N=941)**<br>Estimate [95% CI] |
| --- | --- | --- | --- |
| Age | NS | NS | **0.10 [0.06; 0.14]** |
| Gender (baseline = male) | -0.84 [-2.19; 0.50] | NS | NS |
| Marital status (baseline = married) | NS | NS | **0.74 [0.02; 1.47]** |
| Education | **1.04 [0.31; 1.76]** | **1.68 [1.09; 2.26]** | **1.40 [0.75; 2.05]** |
| Current employment | -0.61 [-2.07; 0.85] | -0.46 [-1.64; 0.73] | 0.29 [-0.80; 1.37] |
| Socio-economic status | 0.80 [-0.10; 1.71] | **1.35 [0.54; 2.15]** | **1.59 [0.81; 2.38]** |
| Nationality (baseline = Lebanese) | NA | NA | **- 1.63 [- 3.04; - 0.21]** |
| Willingness to approach immediate family/relatives for help | NS | 0.75 [-0.87; 2.37] | 0.16 [- 1.29; 1.62] |
| Health coverage | **2.31 [0.94; 3.69]** | **4.55 [3.38; 5.72]** | **2.82 [1.74; 3.89]** |
| Barriers to care | **-0.67 [-0.87; -0.46]** | **-0.68 [-0.86; -0.50]** | **-0.28 [-0.45; -0.11]** |
| Main provider | **2.08 [1.38; 2.79]** | **2.85 [2.20; 3.51]** | **1.63 [1.02; 2.23]** |
| Visits to private clinics | **0.65 [0.09; 1.21]** | 0.06 [-0.30; 0.42] | - 0.11 [- 0.43; 0.21] |
| Visits to PHC centres | -0.43 [-0.89; 0.03] | -0.10 [-0.44; 0.23] | - 0.18 [- 0.48; 0.12] |
| Visits to pharmacy (consultations) | **0.75 [0.31; 1.19]** | **0.83 [0.45; 1.21]** | **0.67 [0.32; 1.01]** |
| Visits to Hospital ER | -0.20 [-1.41; 1.02] | NS | - 0.15 [- 1.13; 0.82] |
| Hospital admissions | 0.33 [-0.93; 1.59] | 0.62 [-0.19; 1.43] | 0.77 [0.18; 1.72] |

NA = Not applicable; NS = Not significant at the bivariate level / not included in the model

Across Lebanese participants, positive perceptions of the care model were significantly associated with participants having health coverage and higher levels of educational attainment. This relationship remains significant even when accounting for barriers to care seeking. Further, perceptions of the care model differed according to the type of health care provider that participants sought care from: the total care model score was highest for those seeking care from specialists, and gradually lower for the other categories (family physicians, general practitioners and lowest for pharmacists). Consultation visits to private clinics and to pharmacies are also associated with increases of the care score.

For Syrian participants, positive perceptions of the care model were also associated with participants having health coverage, higher levels of educational attainment and with being part of a higher socio-economic group. The effect of health coverage was higher for this group compared to Lebanese participants. Barriers to care remained a negative influence and differences between the types of care providers were similar among this group as for the Lebanese one. Visits to private clinics had no bearing on perceptions of the care model, however, visits to pharmacies were still associated with increases in the care model score.

### Section 3: Trust in the health system

Participants were asked for their average level of agreement relating to a series of statements corresponding to the Ozawa et al. (2013) framework on trust in the health system.^15^

High levels of agreement were reported in relation to statements on communication with health providers, confidentiality, competence, and honesty. Lebanese respondents ranked the related statements in this same order with corresponding percentages of agreement decreasing from 92.9% (for communication) to 78.2% (for honesty). Syrian refugees scored those statements very high as well, but communication moved to fourth place (with an agreement percentage of 86.4%) after the following: confidentiality (94.3%), competence (91.0%) and honesty (88.5%).

Lower levels of agreements were identified for confidence in the reliability of the health system (67.4% of Lebanese respondents and 77.6% of Syrian respondents), fairness of the system to provide care to disadvantaged and vulnerable groups (57.3% of Lebanese respondents and 72.5% of Syrian respondents) and the fidelity of health providers to work beyond self-gain (46.8% of Lebanese respondents and 55.9% of Syrian respondents). Finally, the statement around the overall system trust scored at 78.6% of agreement among the Lebanese group and 84.5% among the Syrian group. (See appendix 7 for full results)

#### Influences on trust in the health system

Limited differences between the trust score of Lebanese and Syrian refugee respondents are evident (see figure 5): the median among both groups are closely aligned and while views of Syrian refugees appear more positive and show less dispersion, it is clear that outliers exist.

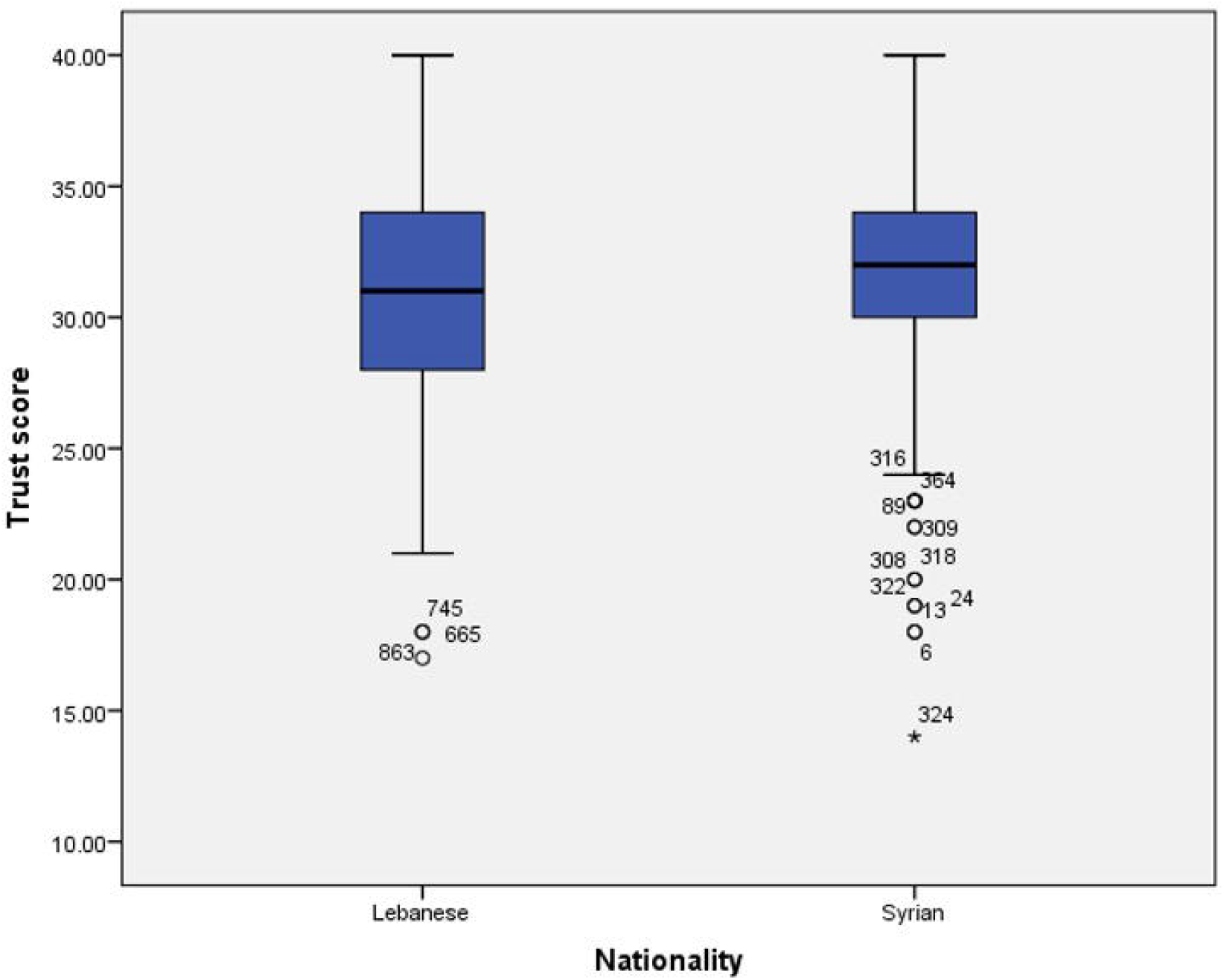

Bivariate analyses suggested multiple variables (most socio-demographic and barriers to care, sources of health seeking, care model score and reported health status) significantly influenced trust in the health system. However, many of those variables were no longer significant in the multivariable analysis (presented in table 5). Importantly, trust in the health system was negatively associated with health status, however this relationship was not statistically significant.

**Table 5.** Multivariable linear regression of trust in healthcare (outcome) and covariates among people living with NCDs in Greater Beirut – Lebanon (2021)

| **Variables** | **Lebanese (n=574)**<br>Estimate [95% CI] | **Syrian (n=367)**<br>Estimate [95% CI] | **Both communities**<br>Estimate [95%CI] |
| --- | --- | --- | --- |
| Age | -0.02 [-0.04; 0.01] | NS | -0.02 [-0.04; 0.01] |
| Education | 0.21 [-0.20; 0.62] | -0.09 [-0.55; 0.38] | 0.20 [-0.11; 0.51] |
| Employment | 0.01 [-0.66; 0.67] | NS | -0.97 [-1.34; -0.60] |
| Nationality | NA | NA | **2.05 [1.41; 2.69]** |
| Health coverage | 0.03 [-0.66; 0.73] | **0.98 [0.14; 1.83]** | 0.47 [-0.05; 0.99] |
| Barriers to care | **-0.32 [-0.43; -0.21]** | -0.08 [-0.20; 0.04] | **-0.26 [-0.34; -0.18]** |
| Main provider | **0.44 [0.08; 0.80]** | **1.16 [0.66; 1.67]** | **0.59 [0.31; 0.88]** |
| Visits to private clinics | 0.08 [-0.20; 0.30] | 0.17 [-0.01; 0.36] | **0.17 [0.02; 0.32]** |
| Visits to PHC centres | -0.08 [-0.31; 0.15] | NS | NS |
| Visits to pharmacy (consultations) | **-0.28 [-0.49; -0.06]** | 0.11 [-0.15; 0.37] | -0.07 [-0.24; 0.09] |
| Visits to hospital ER | 0.20 [-0.40; 0.79] | 0.41 [-0.33; 1.15] | 0.23 [-0.23; 0.69] |
| Hospital admissions | -0.16 [-0.78; 0.45] | 0.08 [-0.59; 0.75] | -0.23 [-0.68; 0.22] |
| Care model score | **0.25 [0.20; 0.29]** | **0.17 [0.14; 0.21]** | **0.22 [0.19; 0.25]** |
| Health Status | NS | 0.01 [-0.55; 0.56] | -0.37 [-0.77; 0.02] |

For Lebanese participants, increases in care model score and decreases in barriers to seeking care are both significantly associated with increases in the trust score. The outcome increases by 0.25 (95% CI = 0.20; 0.29) for each one-unit increase in the care model score and by 0.32 (95% CI = 0.21; 0.43) for each one-unit decrease in the ‘barriers to care’ score. The type of health provider and visits to pharmacy for consultations are also significantly associated with trust in care. Analyses suggest that the trust scores are highest among those seeking care at specialists. In contrast, seeking care from the pharmacy was associated with a decrease in trust. Health coverage had limited bearing on trust, and trust itself was not significantly associated with health status (i.e. disproving the relationship that improved outcomes may bolster trust in the system).

For Syrian refugees, a positive perception of the care model remained significantly associated with higher levels of trust. The type of the main provider was also significantly associated with trust in healthcare with a higher estimate (1.16 with 95%CI = 0.66; 1.67) compared to the Lebanese sub-group. Health coverage remained significantly associated with the outcome at this multivariable level (estimate = 0.98 and 95%CI = 0.14; 1.83) unlike among the Lebanese community. Trust was positively associated with health status, however this relationship was not statistically significant.

## Discussion

Our findings indicate that access to NCD services in the capital of Lebanon is inequitable, with Syrian refugees experiencing more barriers to care seeking compared to host community members. Principal factors that affect health seeking and service accessibility include refugee status, poor socio-economic status and absence of health coverage. Findings indicate that the two communities seek care from different sources: Syrian refugees rely primarily on primary care centres and dispensaries, compared to Lebanese host community members who primarily seek care from specialists. Pharmacies were also identified as an important source of health service provision.

Primary barriers to care seeking were the same among both communities and relate to financial challenges to care access. For Lebanese persons, health coverage, gender and employment were important influences on access to care; for Syrian refugees, socio-economic status only is statistically significantly associated with this. Health status among both communities was poor overall: approx. 5 in 10 Lebanese community members reported being in good health, compared to 4 in 10 for Syrian refugees.

Perceptions of the care model significantly differ by community. Lebanese community members generally have more positive perceptions compared to Syrian refugees. Among Lebanese participants this can be partly attributed to presence of health coverage and level of education, even when accounting for barriers to care seeking. However, for this group of participants, positive perceptions correlated with the type of health care provider accessed: those accessing specialists and private clinics were likeliest to have positive perceptions. For Syrian refugees, health care coverage, level of education and socio-economic status also influenced perceptions of the care model, but barriers to care negatively influenced perceptions. There were no differences in perceptions depending on whether care was sought from private providers.

Trust in the health system was higher among Syrian compared to Lebanese participants, but differences in scores were notable by item. For example, approx. half of Lebanese participants perceived the system as fair in its care provision to disadvantaged and vulnerable groups, compared to almost three quarters of Syrian refugees. Trust was significantly influenced by the care model score and barriers to care seeking.

Evidence on inequitable access to health services is high in LMIC even with the implementation of health reforms and interventions.^23^ Levesque et al. (2013) conceptualised access to care by integrating factors from both supply and demand sides and the context (e.g. urban areas), and summarized them in five concepts: approachability; acceptability; availability and accommodation; affordability; and appropriateness.^24^ Our study findings showed gaps in all those dimensions and validated that reducing inequities in access to care should tackle different elements such as cultural and psychological barriers (affecting acceptability), and financial barriers to care affecting the capacity of people to use health services.

Our findings identified disparities in terms of services provided to different community members by exploring how different communities perceived the NCD care received. Empirical research in other LMIC settings also investigated perceptions and attributes of the care model and explored inequities in the distribution of primary care services. For instance, Pongpirul et al. (2009) reported satisfactory but inequitable features of primary care between different regions in Thailand and therefore identified pitfalls in the PHC policy of the country that need to be addressed.^25^ Our study also highlighted more positive perceptions – likely associated with better care features – within the private sector. Similar findings were reported by a study in Hong Kong where primary care experiences were better when received from private providers,^26^ showing the importance of such approaches to understand the differences in primary care quality of services in the case of multiple providers. In terms of the specialty of providers, a study in Taiwan showed that patients visiting primary care physicians reported better experiences in relation with several domains such as continuity, coordination and comprehensiveness of services, compared to those seeking care from specialists.^27^ These findings from a setting where there is no restriction on physician choice such as Lebanon contradict the findings of our study, suggesting that more support is needed for the primary care workforce in Lebanon to take the lead on providing essential services and have the power to be gatekeepers of the health system.

This survey offered insights on the complexity of trust in healthcare and its determinants. The first observation is that community members identified gaps in different aspects of trust – related to both interpersonal trust and institutional trust, confirming the dynamic and multi-dimensional characteristics of this concept.^28^ Of major importance were the lower scores on the reliability and fairness of the system (elements of institutional trust) and the fidelity of providers to work beyond self-gain (element of interpersonal trust), compared to other dimensions such as competence of providers and confidentiality. This difference in perceptions between domains may relate to general perceptions of commercialization of healthcare in Lebanon,^16^ and confirms that structural reforms in the system towards universal health coverage would contribute to increasing trust of communities in the health system.

Our findings also validated hypotheses from previous research studies on the impact of accessibility of care, primary care features of provided services and the type of providers on people’s trust in healthcare.^17-18^ However, an unexpected observation was the negative association between receiving pharmacist consultations and trust in healthcare, even though this same variable of pharmacist consultations was associated with more positive perceptions of care. A reasonable explanation is that community members view accessing pharmacists as easier compared to physicians but acquire negative perceptions on the health system because of the need to find such alternatives. Therefore, the discussion about the role of different providers within the health system in Lebanon and in other fragile contexts should take into account providers contribution to the delivery of quality services and also the impact on community trust in the system and its implications of their relationship with the system.

### Strengths and limitations

This survey adds to the literature on NCD care delivery and its perceptions from diverse communities in Lebanon, a fragile context. Given the robust sampling methods used, findings are generalisable to other urban contexts in Lebanon, and thus can provide high quality evidence for informing health system strengthening approaches which promote equity and responsiveness to community needs. However, a few limitations can be identified. First, findings cannot be used to extrapolate on care delivery in rural contexts in Lebanon or elsewhere. Second, social desirability bias could have contributed to participants noting higher levels of trust in healthcare among the refugee community despite poorer perceptions of the primary care model among them. Nonetheless, the high similarity in the rank of trust dimensions and in some determinants of trust between the study subgroups suggests a low risk of bias.

## Conclusion

Our study suggests that communities in Lebanon experience and perceive differences in NCD care access and provision. Evidence generated from this paper could guide service delivery reforms and inform how to make the process and targets of NCD service delivery for NCDs in Lebanon and other similar fragile contexts more inclusive and person-centred.

## Data Availability

All data produced in the present study can be made available upon reasonable request to the authors

## Funding and acknowledgment

This research was funded by the National Institute for Health Research (NIHR) (16/136/100lΔNIHR Research Unit on Health in Situations of Fragility – RUHF) using UK aid from the UK Government to support global health research. The views expressed are those of the authors and not necessarily those of the UK National Health Service, the NIHR or the UK Department of Health and Social care. Data collection was conducted by Statistics Lebanon.

## References

1. Vos T, Lim SS, Abbafati C, Abbas KM, Abbasi M, Abbasifard M, Abbasi-Kangevari M, Abbastabar H, Abd-Allah F, Abdelalim A, Abdollahi M. Global burden of 369 diseases and injuries in 204 countries and territories, 1990–2019: a systematic analysis for the Global Burden of Disease Study 2019. The Lancet. 2020 Oct 17;396(10258):1204–22.

2. Lancet T. COVID-19: a new lens for non-communicable diseases. Lancet (London, England). 2020 Sep 5;396(10252):649.

3. World Health Organization. Best buys’ and other recommended interventions for the prevention and control of noncommunicable diseases. Geneva: World Health Organization. 2017.

4. Van Lerberghe W. The world health report 2008: primary health care: now more than ever. World Health Organization; 2008.

5. Ager A, Saleh S, Wurie H, Witter S. Health systems research in fragile settings. Bulletin of the World Health Organization. 2019 Jun 6;97(6):378.

6. Diaconu K, Falconer J, Vidal N, O’May F, Azasi E, Elimian K, Bou-Orm I, Sarb C, Witter S, Ager A. Understanding fragility: implications for global health research and practice. Health policy and planning. 2020 Mar 1;35(2):235–43.

7. Jailobaeva K, Falconer J, Loffreda G, Arakelyan S, Witter S, Ager A. An analysis of policy and funding priorities of global actors regarding noncommunicable disease in low-and middle-income countries. Globalization and health. 2021 Dec;17(1):1–5.

8. Akik C, Asfahani F, Elghossain T, Mesmar S, Rabkin M, El Sadr W, Fouad FM, Ghattas H. Healthcare system responses to non-communicable diseases’ needs of Syrian refugees: The cases of Jordan and Lebanon. Journal of migration and health. 2022 Jan 1;6:100136.

9. Witter S, Zou G, Diaconu K, Senesi RG, Idriss A, Walley J, Wurie HR. Opportunities and challenges for delivering non-communicable disease management and services in fragile and post-conflict settings: perceptions of policy-makers and health providers in Sierra Leone. Conflict and health. 2020 Dec;14(1):1–4.

10. Ajisegiri WS, Abimbola S, Tesema AG, Odusanya OO, Peiris D, Joshi R. The organisation of primary health care service delivery for non-communicable diseases in Nigeria: a case-study analysis. PLOS Global Public Health. 2022 Jul 1;2(7):e0000566.

11. Gupta N, Coates MM, Bekele A, Dupuy R, Fénelon DL, Gage AD, Getachew T, Karmacharya BM, Kwan GF, Lulebo AM, Masiye JK. Availability of equipment and medications for non-communicable diseases and injuries at public first-referral level hospitals: a cross-sectional analysis of service provision assessments in eight low-income countries. BMJ open. 2020 Oct 1;10(10):e038842.

12. Gouda HN, Richards NC, Beaglehole R, Bonita R, Lopez AD. Health information priorities for more effective implementation and monitoring of non-communicable disease programs in low-and middle-income countries: lessons from the Pacific. BMC medicine. 2015 Dec;13(1):1–8.

13. Allen LN, Pullar J, Wickramasinghe KK, Williams J, Roberts N, Mikkelsen B, Varghese C, Townsend N. Evaluation of research on interventions aligned to WHO ‘Best Buys’ for NCDs in low-income and lower-middle-income countries: a systematic review from 1990 to 2015. BMJ global health. 2018 Feb 1;3(1):e000535.

14. Arakelyan S, Jailobaeva K, Dakessian A, Diaconu K, Caperon L, Strang A, Bou-Orm IR, Witter S, Ager A. The role of trust in health-seeking for non-communicable disease services in fragile contexts: A cross-country comparative study. Social Science & Medicine. 2021 Dec 1;291:114473.

15. Ozawa S, Sripad P. How do you measure trust in the health system? A systematic review of the literature. Social science & medicine. 2013 Aug 1;91:10–4.

16. Bou-Orm I, Loffreda G, Diaconu K, Witter S, DeVos P. Political Economy of Non-Communicable Disease (NCD) Prevention and Control in Lebanon: Identifying Challenges and Opportunities for Policy Change and Care Provision Reforms. Under review – Plos One.

17. Zablith N, Diaconu K, Naja F, El Koussa M, Loffreda G, Bou-Orm I, Saleh S. Dynamics of non-communicable disease prevention, diagnosis and control in Lebanon, a fragile setting. Conflict and health. 2021 Dec;15(1):1–3.

18. Zablith N, Diaconu K, Naja F, El Koussa M, Loffreda G, Bou-Orm I, Saleh S. Dynamics of non-communicable disease prevention, diagnosis and control in Lebanon, a fragile setting. Conflict and health. 2021 Dec;15(1):1–3.

19. Bou-Orm I, Adib S. Prevalence and clinical characteristics of diabetes mellitus in Lebanon: a national survey. Eastern Mediterranean Health Journal. 2020 Feb 1;26(2).

20. Shi L, Starfield B, Xu J. Validating the adult primary care assessment tool. Journal of Family Practice. 2001 Feb 1;50(2):161.

21. Comission P. Social Capital: reviewing the concept and it’s policy implications. Canberra, AusInfo. 2003:1–89.

22. Cislaghi B, Cislaghi C. Self-rated health as a valid indicator for health-equity analyses: evidence from the Italian health interview survey. BMC Public Health. 2019 Dec;19(1):1–3.

23. Umeh CA, Feeley FG. Inequitable access to health care by the poor in community-based health insurance programs: a review of studies from low-and middle-income countries. Global Health: science and practice. 2017 Jun 27;5(2):299–314.

24. Levesque JF, Harris MF, Russell G. Patient-centred access to health care: conceptualising access at the interface of health systems and populations. International journal for equity in health. 2013;12(1):1–9.

25. Pongpirul K, Starfield B, Srivanichakorn S, Pannarunothai S. Policy characteristics facilitating primary health care in Thailand: a pilot study in transitional country. International Journal for Equity in Health. 2009 Dec;8(1):1–8.

26. Wong S, Kung K, Griffiths SM, Carthy T, Wong M, Lo SV, Chung VC, Goggins WB, Starfield B. Comparison of primary care experiences among adults in general outpatient clinics and private general practice clinics in Hong Kong. BMC public health. 2010 Dec;10(1):1–1.

27. Tsai J, Shi L, Yu WL, Hung LM, Lebrun LA. Physician specialty and the quality of medical care experiences in the context of the Taiwan national health insurance system. The Journal of the American Board of Family Medicine. 2010;23(3):402–12.

28. Gilson L. Building trust and value in health systems in low- and middle-income countries. Social Science and Medicine. 2005; 61(7):1381–1384.

